# Space-time Classification Index for Assessing COVID-19 Hotspots

**DOI:** 10.1101/2021.11.17.21266461

**Authors:** David Haynes, Chetan Tiwari

## Abstract

**Objectives:** To develop new methods to address problems associated with use of traditional measures of disease surveillance, including prevalence and positivity rates.

**Methods:** We use data from the public New York Times Github repository to develop a space-time classification index of COVID-19 hotspots. The Local Indicator of Spatial Association (LISA) statistic is applied to identify daily clusters of COVID-19 cases, from July 4th to July 19th.

**Results:** The classification index is a spatial and temporal assessment tool that seeks to incorporate temporal trends of the clusters that are “high-high” and “high-low”. Two classifications support the index: severity and temporal duration. We define severity as the number of times a county is statistically significant and temporal duration captures the number of consecutive days a county is a hotspot.

**Conclusions:** The space-time classification index provides a statistically robust measure of the spatial patterns of COVID-19 hotspots. Spatial information is not captured through measures like the positivity rate, which merely divides the number of cases by tests conducted. The index proposed in this paper can guide intervention efforts by classifying counties with six-levels of importance.

## Introduction

COVID-19 is the fastest spreading disease in modern history, infecting over 21 million people and killing over 750,000 people in approximately 9 months. Limiting direct interaction among people through social distancing, use of face masks, and restricted access to retail and public spaces is the most effective way to control spread until vaccinations become widely available. While social-distancing measures yield significant public health benefits, they might also have devastating effects on local economies. The lack of reliable statistical measures for assessing the impacts of COVID-19 at fine geographic scales have contributed to the adoption of “one-size-fits-all” policy measures, such as statewide economic shutdowns (1), that are deemed inadequate in some areas or unnecessary in others. We argue that ignoring local geographic trends in COVID-19 infection can lead to ineffective public health recommendations. Moreover, attempts to understand COVID-19 burdens at fine geographic scales are severely limited because of inconsistencies in populations’ access to testing and data collection and reporting protocols.

The positivity rate of COVID-19, defined as the proportion of tests reported positive, is a commonly used measure for evaluating the severity of COVID-19. However, testing data is questionable due to several challenges, including variability in test processing times; test reliability; lack of name-based tracking when multiple tests (PCR, Antigen, and Antibody) are administered; and deficiencies in the data infrastructure needed to track cases and corresponding tests (2). Current practices for calculating the positivity rate, which simply involves dividing the average number of positive tests reported over a certain time period divided by the average number of tests conducted during that same period, does not guarantee that every person in the numerator (i.e., positive case) has one, and only one, corresponding record in the denominator (i.e., persons tested). Positivity rates are typically computed as 7-or 14-day moving averages for regions containing large populations (i.e., states) to address these issues (3). When positivity rates are computed for places with small population sizes, such as rural areas, inconsistencies in case and testing data are exaggerated and present an inaccurate, and potentially misleading, picture of the disease in those areas. The lack of reliable fine-scale metric for assessing COVID-19 burdens severely impacts the ability of public health practitioners to plan and implement intervention measures. In this paper, we present a spatially-explicit approach for identifying county-level trends based on occurrence and persistence patterns of statistically significant hotspots of localized infection.

## Methods and Results

Local Indicators of Spatial Association (LISA) identifies local, or fine-scale, patterns of spatial association (4,5). LISA computes the correlation between every geographic feature on the map (e.g., county) and its neighbors (e.g., adjacent counties). LISA values range between -1·0 and +1·0, where a value of +1·0 indicates high similarity, a value of -1·0 indicates dissimilarity, and a value of 0 indicates no relationship. Additionally, LISA values are classified into one of four categories: high-high, high-low, low-high, and low-low. These categories characterize the relationship between the feature of interest and its adjacent neighbors. Daily case data obtained from New York Times for the period between July 4^th^ and July 19^th^ (6) are used to identify statistically significant hotspots of high-high and high-low clusters across the United States. We convert these values into a county classification index (Figure 1) that incorporates two dimensions of importance: severity and temporal duration of clustering. Severity is the total number of times a county is classified as a statistically significant cluster in the time-period analyzed, and temporal duration counts the number of continuous days during which a county is statistically significant. The following rules are then used to compute the county classification index:

**Figure 1:**
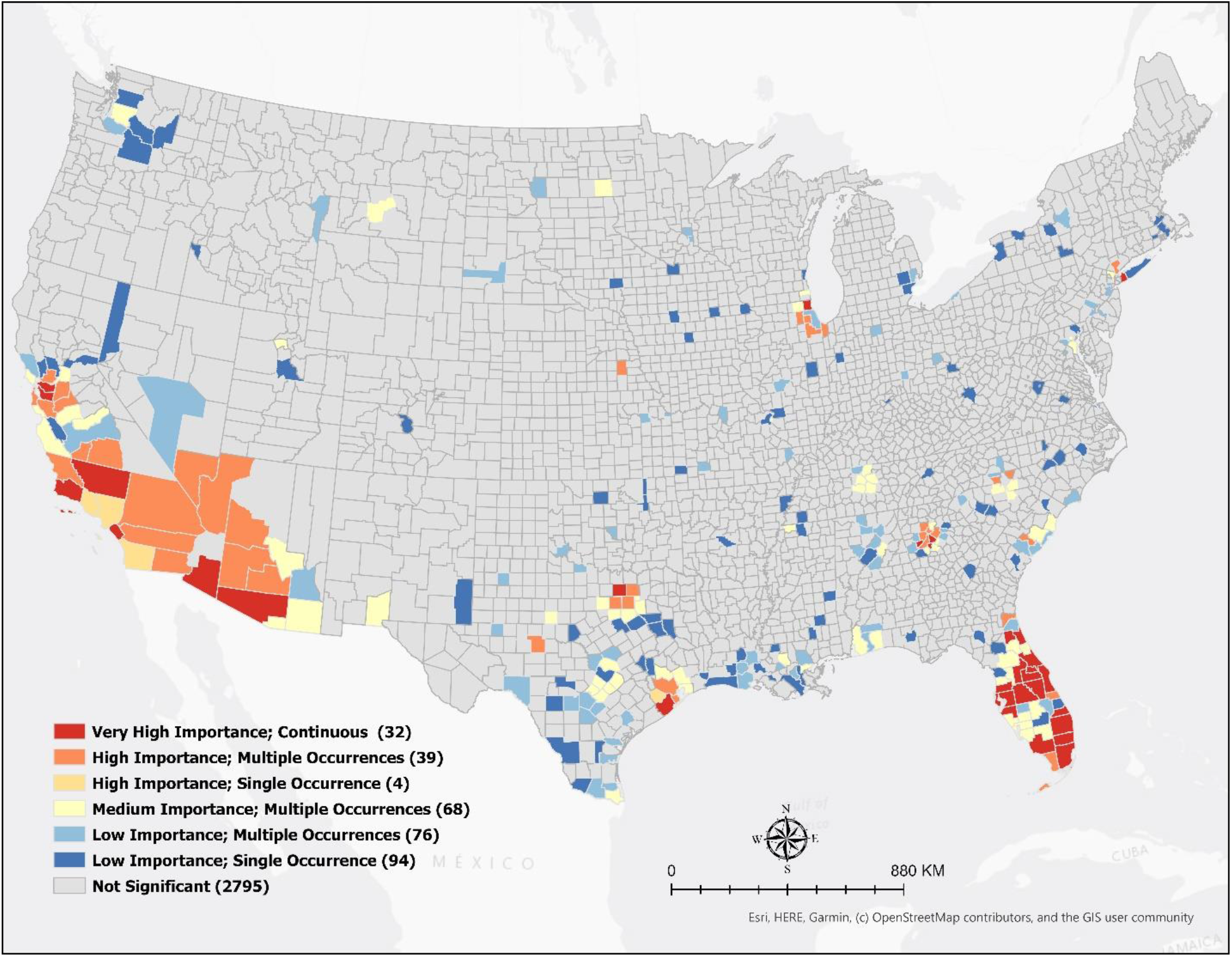
Classified Clusters of COVID-19

### Severity classification

Very High: County is statistically significant every day during the 14-day period

High: County is statistically significant between 10 and 13 days (70% to 99%)

Medium: County is statistically significant between 5 and 9 days (30% to 69%)

Low: County is statistically significant four or fewer days (< 29%)

### Temporal duration classification

Continuous: County is statistically significant every day

Multiple Occurrences: County is statistically significant for at least one day, and this phenomenon repeats throughout the time period

Single Occurrence: County is statistically significant for only a single day

## Discussions and Public Health Implications

The map, Figure 1, reveals hotspots of very high importance in parts of Florida, California, and Texas. These results are consistent with other descriptive assessments of COVID-19 trends during this time-period. For example, Florida reported its highest daily case count (15,300) on July 12^th^, 2020. Note that, in comparison to daily reported cases counts, the map in Figure 1 presents results that are statistically significant. Further, the methodology presented in this paper is not limited to the county level analyses and can be applied to aggregated health data at any geographic scale.

The methodology presented in this paper produces a multivariate index with three important characteristics that are not captured by the positivity rate: (1) it can be used at fine geographic scales (e.g. counties), (2) it identifies statistically significant areas of concern, and (3) it describes spatial and temporal dynamics of COVID-19. Counties highlighted in red in Figure 1 (Very High Importance, Continuous duration) indicate statistically significant hotspots that require immediate and sustained intervention. In contrast, counties in blue (Low Importance) indicate hotspots where strategic intervention such as contact tracking may yield significant benefits.

Note that the LISA statistic identifies hotspots by evaluating geographic features and their neighboring features. Although this statistic does not provide explicit information on the number of cases within each hotspot, we argue that a hotspot with very high or high importance needs immediate attention regardless of the number of cases in that area given high levels of mobility and COVID-19 infectivity. Further details along with interactive maps that can be produced for any time period or interval are available at https://smartcommunityhealth.github.io/solap-template/

## Data Availability

All data produced in the present study are available upon reasonable request to the authors.

https://smartcommunityhealth.github.io/solap-template/

## Data Availability

All data produced in the present study are available upon reasonable request to the authors.

https://smartcommunityhealth.github.io/solap-template/

## Notes

### Competing Interest Statement

The authors have declared no competing interest.

### Funding Statement

This study did not receive any funding

